# Pooled RNA: extraction free testing of saliva for SARS-CoV-2 detection

**DOI:** 10.1101/2022.01.16.22269390

**Authors:** Orchid M. Allicock, Devyn Yolda-Carr, John A. Todd, Anne L. Wyllie

**Affiliations:** Department of Epidemiology of Microbial Diseases, Yale School of Public Health, New Haven, CT 06510, USA; Flambeau Diagnostics, Madison WI 53719, USA

**Author notes:** These authors contributed equally to this article (Orchid M. Allicock, Devyn Yolda-Carr).

**Keywords:** SARS-CoV-2, pooling, real-time RT-PCR, surveillance testing

## Abstract

The key to limiting SARS-CoV-2 spread is to identify virus-infected individuals (both symptomatic and asymptomatic) and isolate them from the general population. Hence, routine weekly testing for SARS-CoV-2 in all asymptomatic (capturing both infected and non-infected) individuals is considered critical in situations where a large number of individuals congregate such as schools, prisons, aged care facilities and industrial workplaces. Such testing is hampered by operational issues such as cost, test availability, access to healthcare workers and throughput. We developed the SalivaDirect RT-qPCR assay to increase access to SARS-CoV-2 testing via a low-cost, streamlined protocol using self-collected saliva. To expand the single sample testing protocol, we explored multiple extraction-free pooled saliva testing workflows prior to testing with the SalivaDirect assay. A pool size of five, with or without heat inactivation at 65°C for 15 minutes prior to testing resulted in a positive agreement of 98% and 89%, respectively, and an increased Ct value shift of 1.37 and 1.99 as compared to individual testing of the positive clinical saliva specimens. Applying this shift in Ct value to 316 individual, sequentially collected, SARS-CoV-2 positive saliva specimen results reported from six clinical laboratories using the original SalivaDirect assay, 100% of the samples would have been detected (Ct value >45) had they been tested in the 1:5 pool strategy. The availability of multiple pooled testing workflows for laboratories can increase test turnaround time, permitting results in a more actionable time frame while minimizing testing costs and changes to laboratory operational flow.

## Introduction

During the emergence and spread of the SARS-CoV-2 virus in 2020, the majority of testing for the virus was aimed at diagnosing COVID-19 (the disease that it causes) in patients presenting with symptoms characteristic of COVID-19. An infected person may develop COVID-19 disease symptoms 3 to 8 days post infection or may never develop symptoms (asymptomatic) (1-3). However, asymptomatic individuals can be infectious, carrying viral loads high enough to spread the virus to uninfected individuals. Soon thereafter it became clear that to control the spread of SARS-CoV-2 infection, a two-pronged approach must be used in the general population to prevent viral spread from infected asymptomatic individuals to the non-infected population.

The first prong involves utilisation of physical barriers (e.g. face masks) to minimize virus spread via aerosols (4, 5). The second prong involves routine weekly testing for SARS-CoV-2 in all asymptomatic (capturing both infected and non-infected) individuals at high-risk for infection (6, 7). Such testing is considered critical in situations where many individuals congregate such as schools, prisons, aged care facilities and industrial workplaces. Testing strategies rely on obtaining a respiratory tract specimen and an assay for the presence of SARS-CoV-2 antigen or genome.

It is generally considered that molecular tests for viral genome are more sensitive than the antigen tests; however, they can be costly, can take days to return results and can be hard to scale for large population testing. Furthermore, for tests requiring a swab-based respiratory tract specimen, these can be uncomfortable and difficult to obtain, especially under weekly self-collected specimen protocols, deterring individuals from participating in testing (8). Early in the pandemic response, saliva emerged as an alternative specimen for SARS-CoV-2 testing and by 2021 it became apparent that a self-collected specimen could obviate the disadvantages of respiratory tract swab specimens. Importantly, the clinical sensitivities for SARS-CoV-2 detection were similar between respiratory tract swab and saliva specimens(9).

To address some of the limitations of testing, SalivaDirect™ was developed as an open-source protocol wherein clinical laboratories could adopt a streamlined, easy-to-use, inexpensive, scalable and flexible genomic (RT-qPCR) assay method for SARS-CoV-2 detection (10). Importantly the assay is based upon a simple self or observed saliva collection protocol. The SalivaDirect™ assay was developed to simplify testing individual saliva specimens; however, with the momentum around testing large-scale asymptomatic populations (e.g. school students, faculty and staff) where SARS-CoV-2 prevalence is low, in a cost-effective manner, a more scalable protocol is required for sustainable testing programs. We therefore investigated higher throughput protocols, wherein saliva specimens are pooled prior to testing with RT-qPCR. These pooled testing approaches were evaluated for the clinical sensitivity of SARS-CoV-2 detection.

## Methods

### Ethics statement

The use of de-identified saliva specimens from healthy or asymptomatic individuals was approved by the Institutional Review Board of the Yale Human Research Protection Program (Protocol ID. 2000028394) (11). Study participants were informed in writing about the purpose and procedure of the study, and consented to study participation through the act of providing the saliva sample; the requirement for written informed consent was waived by the Institutional Review Board. Additionally, the Institutional Review Board of the Yale Human Research Protection Program determined that the use of de-identified, remnant COVID-19-positive clinical samples obtained from laboratory partners for the RT-qPCR testing conducted in this study is not research involving human subjects (Protocol ID: 2000028599).

### Sample Collection

All de-identified saliva samples used in the current study were collected unsupplemented into simple laboratory plastic tubes per the SalivaDirect protocol (12). All samples were tested with the SalivaDirect assay(13) in our research laboratory to confirm SARS-CoV-2 status. Samples were stored at -80°C until further analysis.

### Pooled Sample Testing

To understand the effect of sample dilution by pooling, on clinical sensitivity, a total of 20 saliva specimens which previously tested positive with the modified CDC assay RT-qPCR assay, with resulting cycle threshold (Ct) values between 22.98 and 39.43, were diluted 1:5, 1:10, and 1:20 with negative saliva specimens from healthcare workers(14). Undiluted specimens and pools were tested with the standard SalivaDirect RT-qPCR assay.

After identifying the optimal pool size, we performed an initial workflow evaluation using 5 different pooled samples, each composed of a single SARS-CoV-2-positive saliva sample pooled with 4 SARS-CoV-2-negative saliva samples, in a 1:5 dilution. To confirm our initial workflow findings and assess the sensitivity of viral detection when pooling, 20 additional pools (1 positive with 4 negative saliva specimens) were prepared and tested using the five different workflows.

The five different saliva pooling workflows investigated in both the initial and confirmation studies are depicted in **Figure 1**. All saliva samples were thawed on ice prior to testing and all samples were tested in duplicate. For workflows A and C (**Figure 1a**), 50 μl of each sample, (including the SARS-CoV-2-positive saliva) were pooled to 250 μL total volume, followed by vigorous vortexing. For workflow A, 50 μl of the pooled sample was tested following the standard SalivaDirect protocol(10). For workflow C the remaining sample was treated with 10 μl of proteinase K then heat inactivated before testing directly without further treatment in the SalivaDirect RT-qPCR assay. Workflows B, D and E (**Figure 1b**) involved incubating individual non-pooled samples at 65ºC for 15 minutes before combining 50 μl of each sample into pools of 5 pre-treated samples. For workflow B, 50 μl of the pool of pre-treated samples was tested through the standard SalivaDirect protocol. For workflow E, 10 μl of each of these pre-treated pools was removed and tested with the SalivaDirect RT-qPCR assay without proteinase K treatment. Finally, for workflow D, 10 μl of proteinase K was added to the remaining volume of the pre-treated pool then heat inactivated before testing in the SalivaDirect RT-qPCR assay.

**Figure 1.**
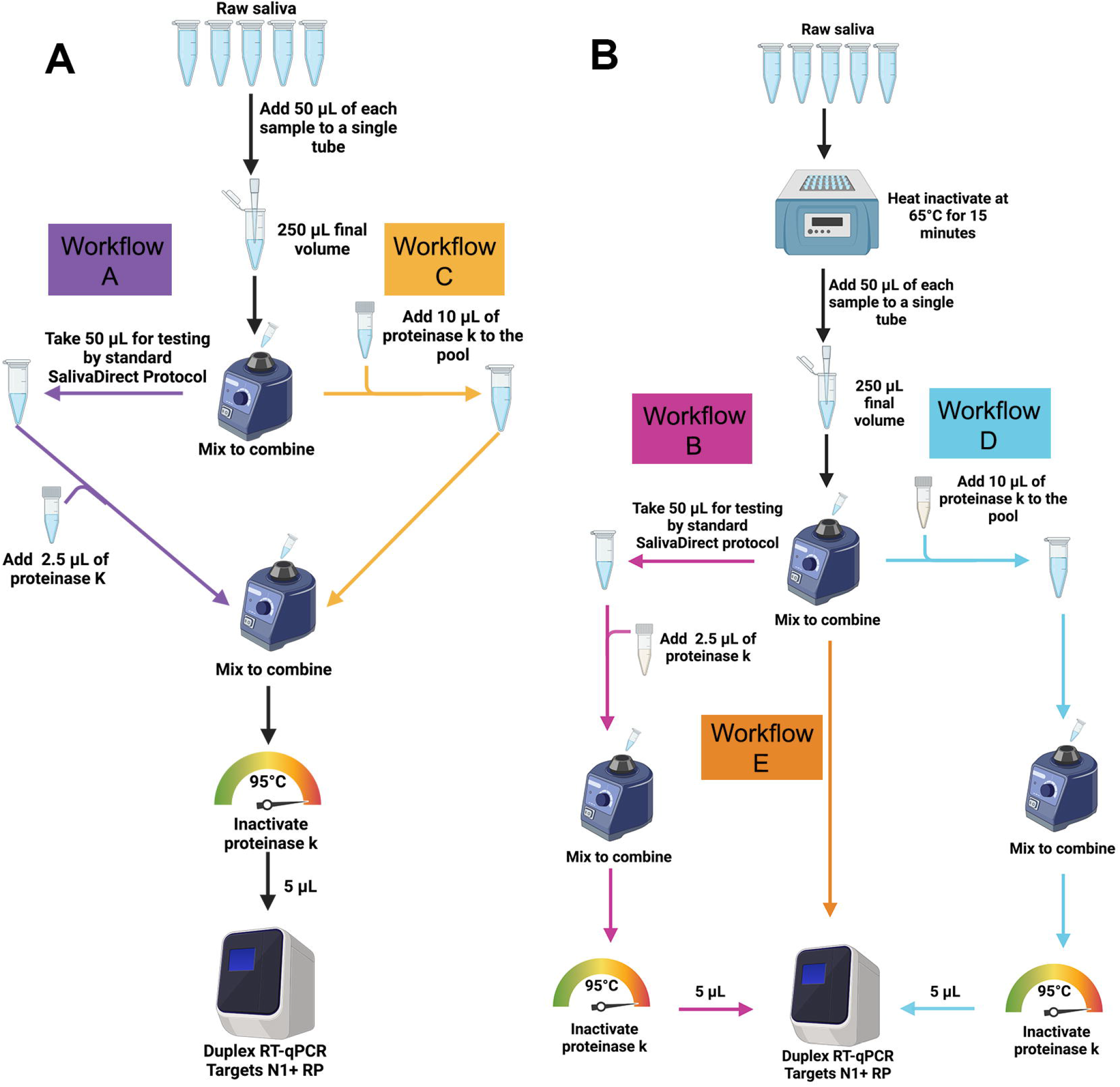
The SalivaDirect™ pooled testing workflows evaluated in the study. (A) For workflows A (purple) and C (gold), 50 μl of SARS-CoV-2-positive saliva and 200 μL total volume of four SARS-CoV-2-negative saliva samples (50 μl each) were pooled and vigorously vortexed to mix. For workflow A, 50 μl of the pooled sample was tested following the standard SalivaDirect protocol. For workflow C the remaining sample was treated with 10 μl of proteinase K then heat inactivated before testing directly without further treatment in the SalivaDirect™ RT-qPCR assay. (B) For workflows B, D and E, individual, non-pooled samples were incubated at 65ºC for 15 minutes before combining 50 μl of each sample into pools of 5 pre-treated samples. For workflow B (pink), 50 μl of the pool of pretreated samples was tested through the standard SalivaDirect protocol. For workflow E (orange), 10 μl of each of these pre-treated pools was removed and tested with the SalivaDirect™ RT-qPCR assay without proteinase K treatment. Finally, for workflow D (blue), 10 μl of proteinase K was added to the remaining volume of the pre-treated pool then heat inactivated before testing in the SalivaDirect™ RT-qPCR assay.

### Assessment of clinical Ct values with pooling

To evaluate the real-world potential loss of sensitivity on clinical samples with pooling, we estimated the average change in Ct value for each pooled testing workflow using the results from the 25-sample workflow confirmation study. We then estimated how the change in Ct value would potentially impact assay sensitivity by applying the ΔCt value (with pooling) to real world SalivaDirect RT-qPCR SARS-CoV-2 testing results. Six SARS-CoV-2 testing sites around the U.S., all testing with the standard SalivaDirect™ protocol, provided sequential testing results (Ct values for positive specimens) during August 2021 and to these values we applied the ΔCt value that we estimated.

### Statistical analyses

The correlation of Ct values between each workflow and the individual positive samples was assessed using the Pearson correlation coefficient and represented graphically with linear regression. The negative RT-PCR of the target gene was set at the Ct value of 45 for the statistical analysis. All statistical analyses were performed using GraphPad Prism version 9 (GraphPad Software, San Diego, CA). For the calculation of percent positive agreement, samples are considered positive at Ct < 45.

## Results

### Pooling sizes and workflow selection

We initially performed a limit of detection range-finding study to determine the impact of sample dilution via pooling with the SalivaDirect RT-qPCR assay on detection sensitivities. As pool size increased the resulting assay Ct values increased as well, generally in a linear manner. The smallest change in Ct values (i.e. loss of assay sensitivity) of pooled versus neat saliva was obtained with 1:5 pooling (1 positive and 4 negative saliva samples). Thus a 1:5 pooling strategy was employed for workflow analysis. Our preliminary results indicated that the SalivaDirect assay was able to detect SARS-CoV-2 in pooled saliva specimens with high virus loads, but additional testing was required to optimize saliva pooling and processing workflows.

Extrapolating from previous work (Watkins et al. 2021), we selected 5 workflows representing different pooling strategies. Initial analyses using 5 pools showed that 4 workflows (A-D) provided similar results for most of the pools (see **Table S1**). Workflow E provided a much larger shift in Ct values for all five pools (5.26) and hence loss of assay sensitivity. As expected, a shift in Ct values (to higher) was noted for all five pooled workflows compared the standard SalivaDirect RT-qPCR assay performed on the undiluted positive sample. For workflows A-E, initial analysis of the differences in Ct values between the pools and individual positive samples resulted in a Ct shift of 2.17 to 3.50. Workflow E was omitted from further evaluations.

### Workflow analysis

Twenty-five SARS-CoV-2 saliva specimens were used for pooling, with each pool including one unique positive specimen and 4 negative specimens, to make 25 contrived pools. The Ct values (obtained at the site of saliva collection) for SARS-CoV-2-positive samples ranged from 22.98 to 39.43. The majority (40 to 45%) of the specimens had Ct values >35 indicating a relatively low concentration of virus, 28 to 32% of the specimens had Ct values <30 indicating a higher concentration of virus (**Table 1, Table S2**).

**Table 1.** Distribution of the Ct values of clinical saliva samples used for pooling.

| Ct range* | No. samples Workflow A<br>(n=22) | No. samples Workflow B - E<br>(n=25) |
| --- | --- | --- |
| 20.0-29.9 | 7 (32%) | 7 (28%) |
| 30.0-34.9 | 5 (23%) | 8 (32%) |
| 35.0-40.0** | 10 (45%) | 10 (40%) |

\*samples <40 Ct are considered positive for SARS-CoV-2
\*\*mean Ct for the CFX96 Touch RT-qPCR instrument when determining LOD for analytical sensitivity using this set of reagents was 36.7

We assessed the sensitivity of each workflow by comparing to see which workflow had the smallest shift in Ct values between the undiluted sample processed with SalivaDirect RT-qPCR assay and the workflow in question, and by comparing which workflow had the smallest number of pools dropping below the sensitivity threshold (between Ct 40 to 45). When compared to undiluted samples processed with the standard SalivaDirect assay, Workflows A and B provided the highest sensitivity (**Figure 2**). Workflows A and B resulted in 3 and 1 pool(s) with Ct increases to 40 and 45, respectively. In contrast, workflows C and D demonstrated the lowest in clinical sensitivity, with loss in detection in 8/25 and 10/25 pools processed by these workflows respectively.

**Figure 2.**
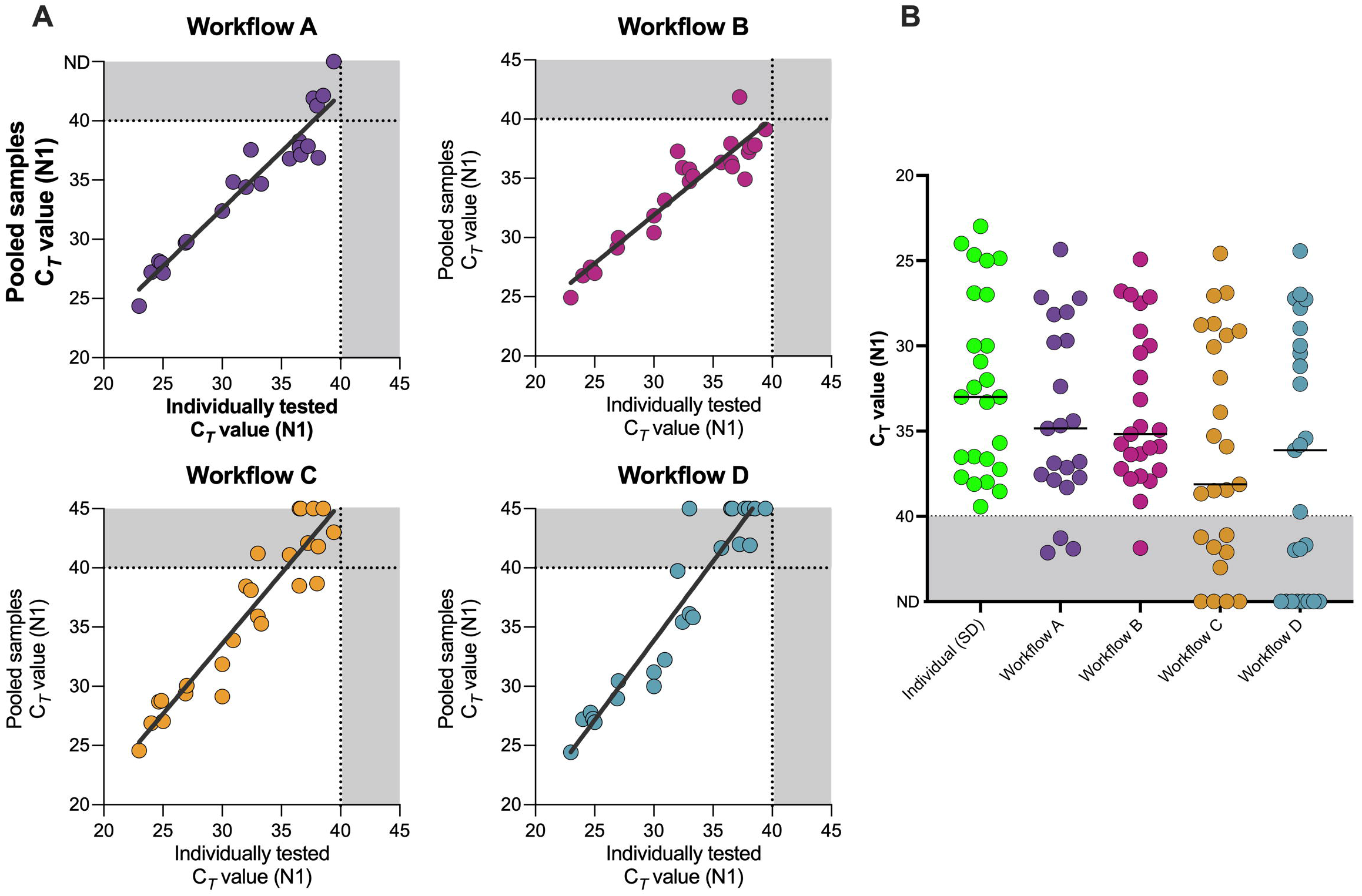
SARS-CoV-2 N1 gene detection with individual saliva-based RNA-extraction-free RT-qPCR testing versus pooled sample testing using Workflows A-D. A) N1 detection of one positive sample pooled and tested with equal volumes of 4 negative samples correlated to the Ct value obtained when samples were tested individually. B). Ct values obtained from each sample tested individually and when combined with 4 negative samples and tested with each of the workflows. The N1 Ct cutoff for classifying individual samples as positive is 40 (as indicated by the grey area under the horizontal dashed line. No cutoff was set for the pooled samples. B) line -median values

Workflow A resulted in a positive agreement of 88.6% (86.4% and 91.0% for the individual replicates), compared to the individual testing results using the standard SalivaDirect protocol. Workflow B resulted in a 98% positive agreement (100% and 96% for the replicates), compared to the individual testing results using the standard SalivaDirect protocol (**Figure 2a**). The positive agreement for workflows C and D were less, with averages of 76% and 62%, respectively.

A theoretical Ct shift of Log_2_(n) can be estimated for most RT-qPCR tests due to the dilution of positive samples when pooled with negative samples. This means that for pools of 5, a Ct value shift of 2.3 would be expected. The Ct shift observed for Workflows A and B were below this expected value, with Ct value shift of 1.99 and 1.37 respectively, confirming a slight loss of assay sensitivity. Workflows C demonstrated the worst Ct value shift of 2.81.

### Impact of pooling on clinical sensitivity

To determine the pragmatic loss of clinical sensitivity with pooling before performing the SalivaDirect RT-qPCR assay with workflows A-D, we queried six SalivaDirect CLIA laboratories across the United States for the Ct values obtained from sequential testing of saliva samples for SARS-CoV-2. These values and the breakdown of samples per site are presented in **Figure 3** (raw data available in **Table S3**). The average Ct value for the six labs was 28.0. There was no statistical difference in Ct values across the labs. Out of a total of 613 determinations across the labs, only 16 samples (2.6%) had Ct values between 38-40. Considering the calculated (from the confirmation study) worst case Ct shift in pooling workflows A and B of 1.99, and if all of the 613 determinations had been made using workflow A or B, these 16 samples would have shifted Ct to between 40 and 42. While considered negative using the individual testing workflow, these 2.6% of samples would fall into a grey zone of 40-45 Ct values.

**Figure 3.**
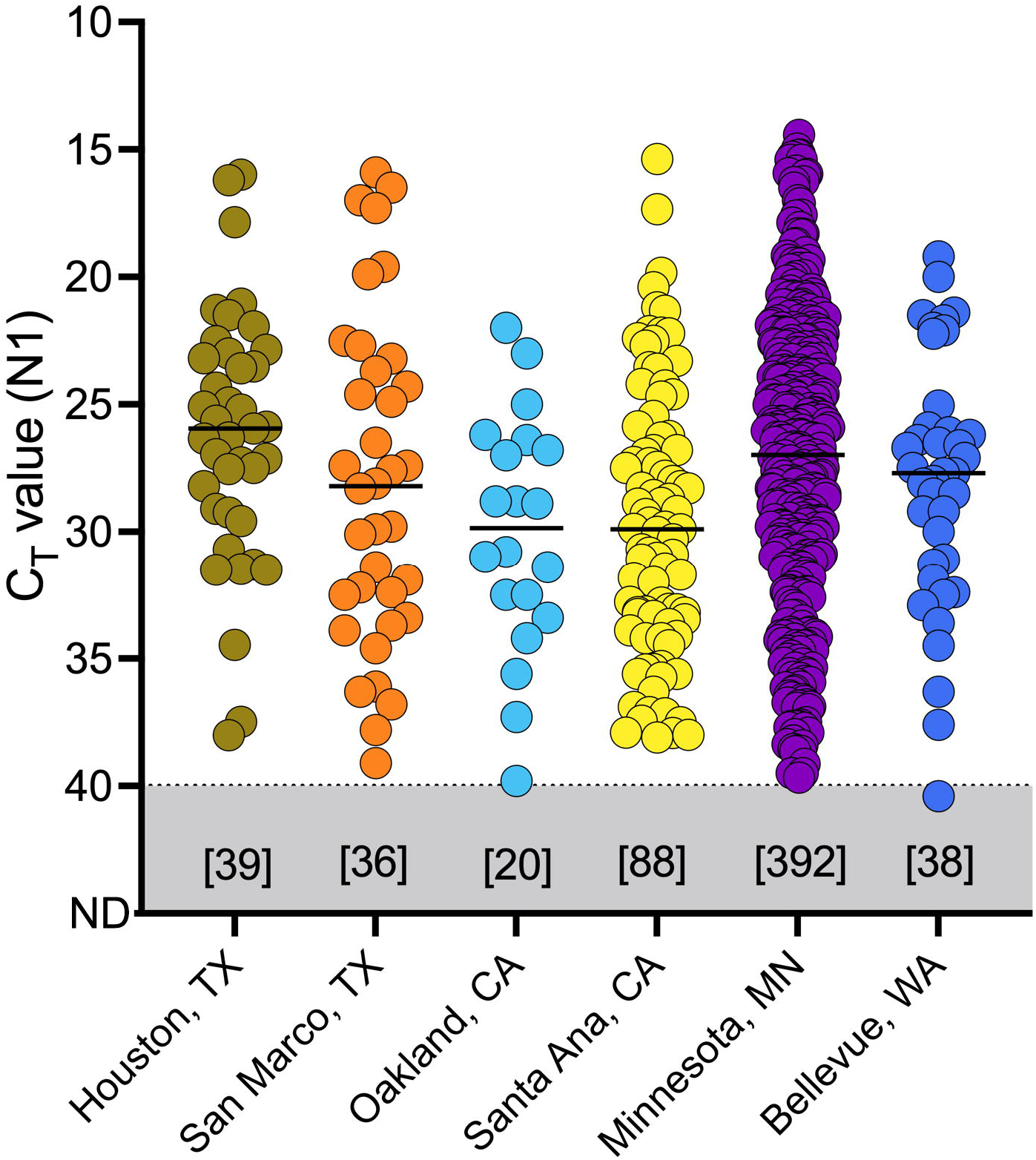
Detection of SARS-CoV-2 N1 gene persists when Ct value shift from workflows A is applied to data from six clinical laboratories across the US in July 2021. Each dot represents the clinical samples. The black line indicates the median value of the samples, and the number of samples processed at each site in square brackets above the location of the laboratory.

## Discussion

Widespread surveillance of asymptomatic individuals is one of the main methods of controlling the spread of SARS-CoV-2. The pooling of samples before testing is a resource-saving approach to increase testing capacity, especially for surveillance in a population with a low infection rate (14), such as travellers, school populations and employees of large organisations. Additionally, testing these members of the community serve as a proxy to the broader community, perhaps identifying larger outbreaks through family members and their associated activities. As saliva is easy to collect from a large number of people, pooling strategies are thus a natural extension to surveillance programs. While pooling saliva does impact assay sensitivity and potentially decrease virus detection, the actual impact appears to be minimal.

In the current study, we demonstrated that weakly positive samples (Ct values of 38 to 40) may be missed when testing pools of larger sizes (pools > 5) when compared to testing samples individually. However, on the basis of the calculated relative sensitivity loss resulting from pooling, we looked at six datasets comprising 613 SARS-CoV-2 positive samples from across the U.S. Using such real world data we found that pooling saliva in groups of 5 samples prior to testing is expected to have minimal impact on clinical sensitivity. Based upon the lab reported Ct values only 2.6% of these samples would have shifted into a Ct 40-45 grey zone using the proposed SalivaDirect pooling workflows A or B. Importantly, if these samples were pooled, none would have become undetectable. It is advised that any sample pool resulting in Ct values between 40-45 should be retested individually using the standard SalivaDirect protocol.

Surveillance programs for SARS-CoV-2 genomic testing in low prevalence populations must be operationally pragmatic. First, they need to be cost-effective. Pooled testing significantly reduces reagent costs, lab personnel cost, and lab resource needs. Second, these programs need to be easy to implement. Self-collection of a simple saliva specimen obviates the need for healthcare workers to collect specimens and the associated personal protective equipment. Third, programs should utilize existing resources for sample collection. Self-collection of saliva can be performed anywhere and the resulting specimen can be deposited at a drop site location (e.g. school or workplace entrance) such that specimens from thousands of participants are collected in a parallel manner. Pooling of specimens once received in the laboratory for testing should fit into established laboratory accessioning and pre-analytic workflows. Finally, the end test results must provide acceptable clinical sensitivities and specificities. We have shown that a saliva-based RNA-extraction-free pooled (1:5) testing strategy results in detection of 97.4-100% SARS-CoV-2-positive samples, as compared to individual testing. Large pooled testing programs have already demonstrated the efficacy of pooled saliva testing for helping to keep schools safely open (15, 16), with pooled samples having a similar sensitivity to the molecular testing of individual samples, in terms of both qualitative and quantitative (comparable Ct values between pooled and individual samples) measures.

Throughout the pandemic, clinical laboratories have been hesitant to implement pooled sample testing (17) due to: 1) stringent workflows which do not fit within existing laboratory operations, 2) a lack of clear guidance on how to implement such methods and 3) the perception that clinical sensitivity of the assay will be lost with pooling. The methods we propose in the current study demonstrate minimal impact on assay sensitivity with 5 samples per pool, and are straightforward extensions of a simple SARS-CoV-2 testing method which can be easily conducted manually, without requiring additional investment. SalivaDirect is a flexible extraction-free platform for RT-qPCR testing. For ease of implementation and safety of lab personnel, multiple workflows (18) were developed for the testing of individual samples. We sought to extend this level of flexibility for labs seeking to offer pooled testing. We demonstrated that workflows A and B provide the best assay sensitivity, with B providing a heat pre-treatment step for labs who require it by local Environmental Health and Safety guidelines. Consequently, workflows A and B were selected as the proposed approaches for pooling of the SalivaDirect test.

Overall, surveillance testing is not generally easy, requiring a pivot by traditionally clinical diagnostic labs, especially when scalable protocols do not exist. Thus, when a decrease in positive cases are observed, there is a psychological and practical desire to decrease testing. However, these dips in COVID-19 cases can lead to a decrease in pandemic preventative measures, which inevitably leads to disease resurgence. Additionally, with the introduction of different variants of concern, the need for affordable and sustainable mass testing strategies only becomes more urgent. Our findings suggest that combining saliva with a practical pooling protocol will enable easier SARS-CoV-2 surveillance testing, especially in resource-limited settings. Such pooled testing has the potential to significantly reduce the overall number of tests and associated costs. This would in turn operationally permit an increased frequency of testing, meaning an increased likelihood of detecting individuals earlier in their infection. This approach should allow broader screening in schools and workplaces for SARS-CoV-2 testing and importantly lay the foundation for managing future upper respiratory infection mediated pandemics.

## Data Availability

All data produced in the present work are contained in the manuscript

## Data Availability

Data from this study is available in the supplemental information.

## Author contributions

A.L.W. conceived the study and developed the study protocol. D.A.Y-C. executed the study. A.L.W. co-ordinated external laboratory data. O.M.A. and A.L.W. analyzed the data. O.M.A. assisted with the design of the statistical analysis. J.A.T., D.A.Y-C, O.M.A., and A.L.W. wrote and edited the manuscript.

## Acknowledgements

We thank our external laboratory partners for their time and cooperation to make this valuable extension of the SalivaDirect assay possible for other laboratories around the country. This work was funded by Flambeau Dx (A.L.W) and Fast Grant from Emergent Ventures at the Mercatus Center at George Mason University (A.L.W).

